# Computational Use of Patient–Provider Secure Messaging Data to Achieve Better Clinical Efficiency and Quality of Communication: A Systematic Review

**DOI:** 10.1101/2025.05.19.25327936

**Authors:** Yawen Guo, Di Hu, Yiliang Zhou, Tianchu Lyu, Kai Zheng

## Abstract

Secure messaging (SM) between patients and providers has seen increasing adoption over the past decade, prompting development of computational methods to enable automation and research to enhance clinical efficiency and quality of communication. Through a systematic review, we examined the extant literature to investigate how previous studies had applied computational analyses to SM data. After screening 1,374 papers, we identified 19 relevant studies published between 2017 and 2024, most of which focused on applications for streamlining clinical workflows, facilitating early disease detection, supporting personalized decision making, and enhancing patient health literacy. Among the computational methods used, BERT was consistently shown to deliver best performance. However, all existing studies were constrained by small-size datasets and limited healthcare settings, leading to inadequate validation and poor generalizability. The results of this review highlight key research gaps, particularly the need for more robust computational approaches that ensure scalability, fairness, and clinical applicability.

## Introduction

The Health Information Technology for Economic and Clinical Health (HITECH) Act was enacted in 2009 to promote the meaningful use of health information technology by healthcare providers^1^. Meaningful use refers to the utilization of certified electronic health records (EHR) to improve the quality of care, engage patients, and ensure the security of patient information. It includes a specific requirement for eligible providers to use secure electronic messaging to communicate with patients to extend access to care, improve transparency, and safeguard patient confidentiality^2^.

Secure messaging (SM), as defined by the Centers for Medicare and Medicaid Services, refers to “any electronic communication between a provider and patient that ensures only those parties can access the communication”^3^. It has become a widely adopted feature in EHR systems and has supported various functionalities such as appointment scheduling, medication refill, lab notification, and delivery of web-based interventions^4,5^. By facilitating communication between patients and providers, SM has become an indispensable tool in modern healthcare organizations to enhance patient empowerment and satisfaction. However, overuse of SM has also led to a growing concern, particularly during and after the COVID-19 pandemic, for its role in increasing the burden on providers, contributing to their burn-out^6–8^. To improve the efficiency in processing large volumes of patient-initiated secure messages, artificial intelligence (AI)-based tools have been developed to help manage message triage and response, accompanied by the introduction of new billing policies for e-visits and secure messaging^9–11^.

Previous reviews on SM have explored its integration into healthcare systems and its impact on communication efficiency and quality. For example, reviews conducted by McGeady et al., Wallwiener et al., Goldzweig et al.—and a few others focusing on specific disease topics—have highlighted the evolution of electronic communication tools, assessing their benefits such as improved healthcare delivery and patient satisfaction, while also addressing challenges such as confidentiality and legal issues^12–15^. A comprehensive understanding of the research literature on SM is crucial for identifying knowledge gaps and informing the development of future tools to enhance clinical workflow, reduce clinician burden, and improve patient care^16–18^.

While there have been studies devoted to applying computational methods to real-world SM data, no systematic review exists that synthesizes these efforts to provide a better understanding on how they contribute to advancing patient–provider communication, clinical decision making, and clinical and health services research. The systematic review study reported in this paper addresses this gap by examining existing models and tools developed from real-world SM data. During the review, we focused on classifying the study objectives, methodologies, data sources and scope, evaluation metrics, and clinical applications of existing research; in addition to providing a structured synthesis of applications built on real-world SM data and identifying key trends, limitations, and directions for future research.

## Methods

We conducted a systematic literature search across databases including PubMed, Scopus, IEEE Xplore, ACM Digital Library, Cochrane CENTRAL, CINAHL, and Web of Science. The search strategy used the following terms: (“**secure messag\***” AND **patient**) OR (“**portal messag\***” AND **patient**) in the title and abstract, restricted to studies published from 2009 onwards. Only articles written in English were included. To ensure the robustness of the search strategy, all searches were repeated and reviewed by a librarian for accuracy. The final search was completed in October, 2024.

We included studies that leveraged real-world SM data between patients and providers to develop computational applications and tools. Since this review study focused on patient–provider communication, studies were excluded if they did not use real-world SM content or if the SM did not occur between patients and providers—for example, communication among caregivers, medical students and faculty members, or across different medical settings (e.g., care transitions). We also excluded studies that examined SM alongside other communication methods and reported results collectively. Additionally, studies were excluded if they solely used existing methods or algorithms for evaluation, without developing new applications or tools based on real-world SM data. We did not include opinion pieces, media articles, commentaries, review articles, abstracts, posters, brief communications, or research letters. The titles and abstracts of the identified articles were independently screened by two authors (YG and DH) using the inclusion and exclusion criteria. Any disagreements were resolved through consensus meetings with a third author (KZ). Articles identified as relevant after the title and abstract screening process then underwent a full-text review to determine final inclusion.

The data extracted from the included papers followed a predetermined structure to systematically capture key elements relevant to our study objective. For computational models and applications developed, we extracted information on the algorithm’s general category (e.g., classification, topic modeling, prediction, named entity recognition), study objective, data source and scope, primary evaluation metrics and performance, comparative evaluation, results, outcomes, and recommendations for future research. Each article was independently coded by two of three researchers (YG, DH, and YZ), all of whom are actively conducting research in clinical informatics and patient–provider digital communication. Discrepancies between reviewers were resolved during consensus meetings or with the input of the third researcher (KZ) as needed.

## Results

We identified 2,151 studies from the databases using the search strategy outlined above. A total of 1,374 duplicate references were identified using Covidence software, and an additional 29 were removed manually. We then screened the titles and abstracts of the remaining 748 studies, excluding 358 that did not meet the inclusion criteria, with full text screening leading to the removal of an additional 391 references. A total of 19 studies were included in the final review. Figure 1 provides an overview of the literature search and screening process.

**Figure 1.**
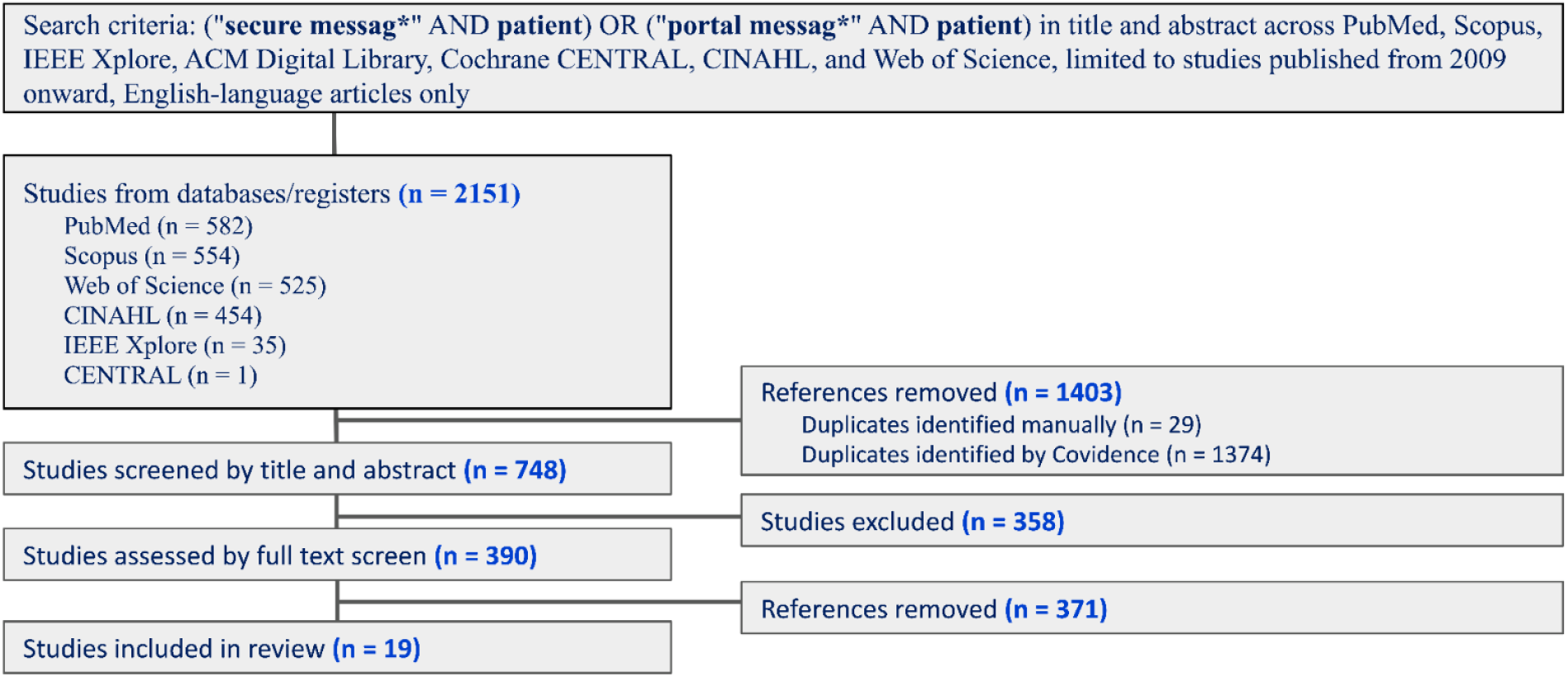
Search strategy and PRISMA flow diagram for the selection of literature

We identified 19 studies focusing on the computational use of patient–provider SM data to develop various applications and tools, published between 2017 and 2024. Most of these studies were published in leading medical informatics venues including Journal of the American Medical Informatics Association (JAMIA) and Journal of Medical Internet Research (JMIR). The studies developed a diverse range of computational approaches to address challenges in patient–provider SM communication through patient portal. Text classification models were the most prevalent (n = 15, 78.9.3%), utilizing methods ranging from traditional ML models, such as logistic regression (LR), support vector machines (SVM), and random forest (RF), to deep learning (DL) models, particularly BERT and its variants^19–22^. Ensemble approaches integrating multiple natural language processing (NLP) techniques were also employed to improve classification accuracy and contextual understanding, such as fusion models combining convolutional neural networks (CNNs) with transformer-based architectures^22,23^. Beyond classification tasks, the studies explored predictive modeling, unsupervised learning, and word embedding optimization to develop computational applications for SM data. One study introduced an automated readability formula using a novel NLP-based approach to predict text difficulty, outperforming traditional Flesch-Kincaid readability scores in assessing message comprehensibility^24^. A study on health literacy profiling developed and validated computational linguistic models to assess patient literacy levels, analyzing 283,216 SMs from diabetes patients to demonstrate that automated literacy profiles could outperform traditional health literacy assessments^25^. A tailored word embedding optimization model called PK-word2vec was designed to address the challenges of small SM datasets, aiming to enhance NLP applications in clinical settings by improving domain-specific language representation^26^. Additionally, a large-scale study leveraging a dataset of two million SMs applied unsupervised learning to uncover common patient concerns and communication patterns, enabling a data-driven exploration of themes in patient–provider interactions^27^.

### Study Objectives

These computational applications were primarily designed to achieve three major objectives: optimizing clinical workflow and triage processes, reducing clinician burden, and enhancing patient communication and health literacy.

#### Optimizing Clinical Workflow and Message Triage (n=5, 26.3%)

Studies aimed to streamline message triage by automatically prioritizing patient messages and classifying urgent and non-urgent inquiries. Text classification techniques were leveraged to categorize messages based on urgency, clinical relevance, and real-time prioritization^20,21,28^. One study specifically classified patient-initiated EHR messages for COVID-19 triage related to medication requests (e.g., Paxlovid), aiming to reduce response time and improve access to antiviral treatment^29^. Additionally, NLP-based approaches were used to predict message complexity and medical decision-making difficulty, further improving workflow efficiency in clinical settings^30^.

#### Reducing Clinician Burden (n=8, 42.1%)

Several studies sought to alleviate clinician workload by automating repetitive tasks and facilitating efficient decision-making. Machine learning applications were developed to detect hypoglycemia incidents in SMs^31^, depression concerns^19^, and identify proxy-authored messages in oncology settings^32^. Several studies classified patient-initiated EHR messages using similar patterns, grouping them into symptoms, prescriptions, logistical concerns, and general updates^33–35^. Some studies further distinguished between informational, medical, and social needs^33,36^. Despite variations in terminology, these studies shared a focus on organizing messages to improve response efficiency and reduce clinician burden.

#### Enhancing Patient Communication and Health Literacy (n=6, 31.6%)

Another major objective was improving patient–provider interactions and understanding patient needs. Several studies developed NLP-based classifiers to assess patient health literacy and predict the readability of physician messages^24,25,37,38^. Others leveraged topic modeling and named entity recognition to extract key themes from patient messages to better understand patient needs^39^, with one study specifically uncovering social needs such as food and housing insecurity, and transportation needs to ensure timely referrals and interventions^36^.

### Data Scale and Source

The scale of data used in the studies varied significantly. We used a cutoff of 10,000 messages to distinguish between small-scale and large-scale studies. The small-scale studies (n=13, 68.4%) primarily focused on analyzing patient-generated messages within limited clinical contexts, such as decision complexity, communicative health literacy, and provider-patient communication patterns^19,21,23,24,28,30–34,36,38,40^. Smaller datasets allowed for greater depth in qualitative analyses but limited interoperability for broader implementation. Large-scale studies (n=6, 31.6%) incorporated datasets of 10,000 or more messages^20,22,25,26,37,39^. These large-scale approaches enabled the application of sophisticated computational approaches but required automated annotation pipelines. The variation in data scale influenced the methodologies used across studies. Small-scale studies relied more on manual annotations, expert reviews, and traditional ML approaches. Large-scale studies leveraged deep learning architectures, such as BERT-based models, and required automated processing pipelines to handle the volume of data effectively.

### Best Performing Models

Across studies, deep learning-based models, particularly BERT and its variants, consistently outperformed traditional ML approaches in classification and triage tasks^19–21,23,29^. For example, a BERT-based large language model achieved an area under the receiver operating characteristic curve (AUROC) of 0.97 in prioritizing patient messages, demonstrating its potential for clinical deployment^41^. Traditional approaches like logistic regression, support vector machine, and random forest, while less dominant, remained competitive in tasks requiring structured feature extraction such as detecting mental health crises and social needs concerns^31,33,40^.

### Comparison of Model Performance

Studies employed various evaluation metrics based on specific tasks. For message triage, prioritization, and information extraction tasks such as extracting patient social needs, metrics such as AUROC, Precision, Recall, and F1-score were commonly used to assess how well models distinguished between urgent and non-urgent messages^21,23,28–30,33,35^. Health literacy and readability analysis utilized readability scores and human ratings to evaluate the effectiveness of NLP models in predicting patient understandability^20,24,25,37,38^. Clinical decision complexity and mental health-related studies relied on classification accuracy, inter-rater agreement, sensitivity, and specificity, ensuring models could reliably distinguish between different levels of clinical significance^19,30,31^. The diversity in evaluation metrics reveals that model evaluation must be tailored to the specific clinical applications, balancing precision, recall, and interpretability to improve real world implementation in patient care.

**Table 1.**
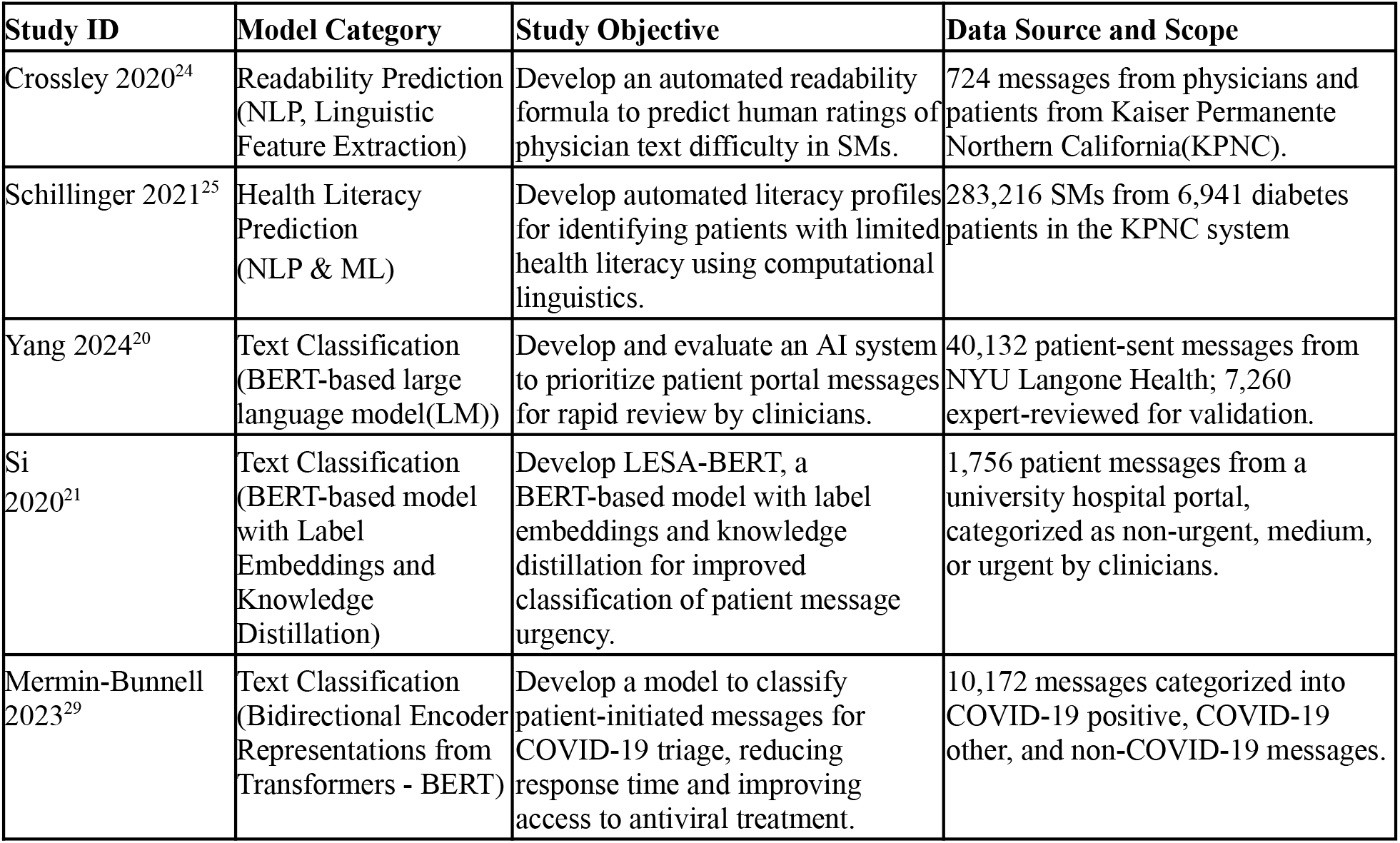
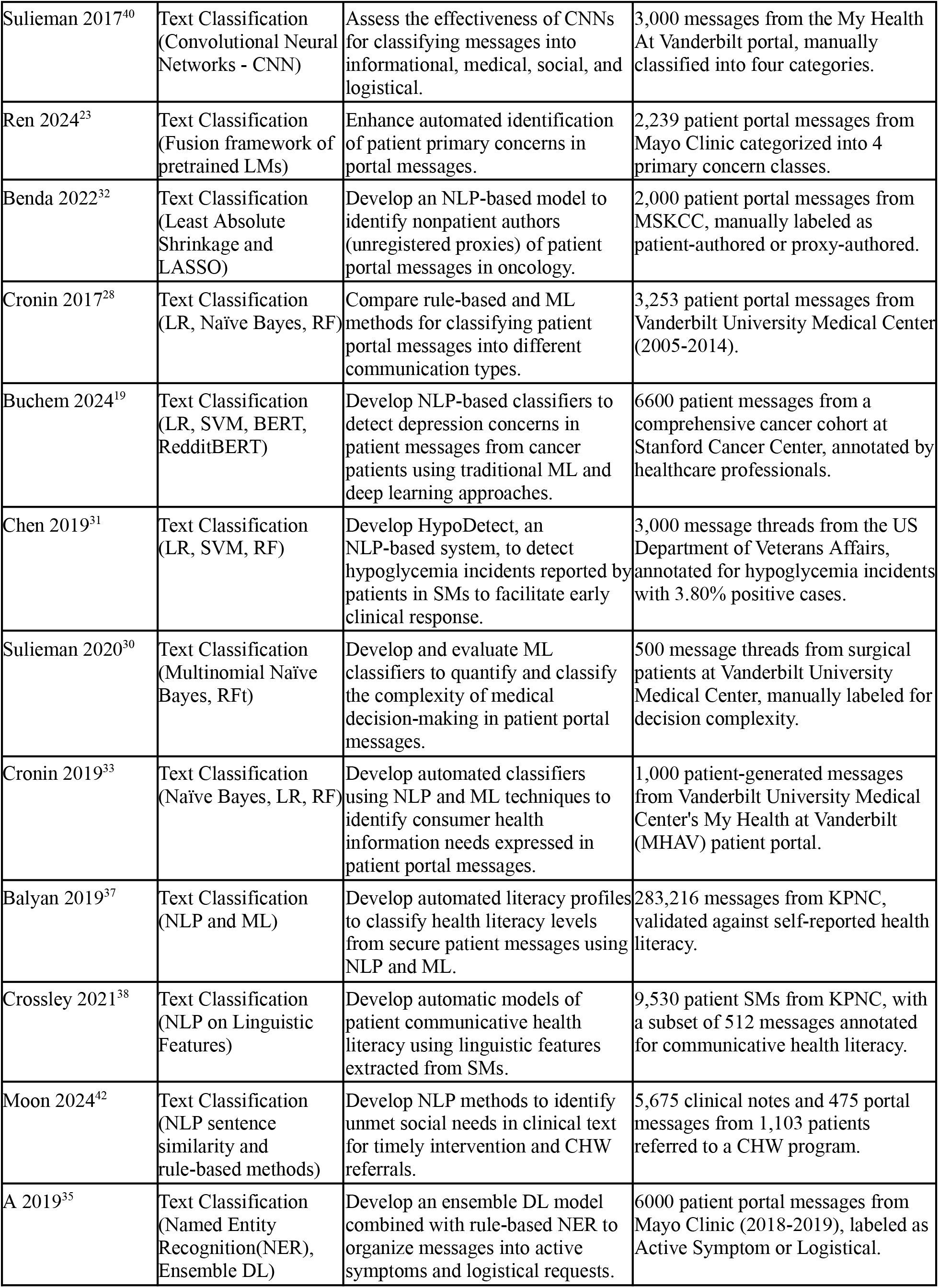
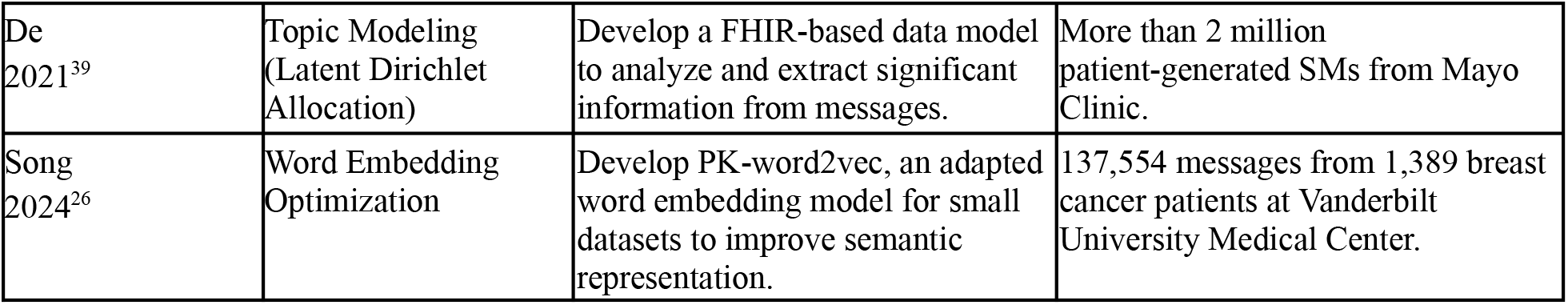
Study Characteristics

## Discussion

Previous studies on patient portals and SM systems have examined system usability, SM communication efficiency, and provider workload. While early evaluations highlighted responsiveness of these systems^43^, later analyses identified barriers such as limited message access, delays in response times^44^, and burdens on clinicians^45^. The COVID-19 quarantine policy and the recent shift in e-visit billing policies have significantly influenced the SM usage and communication patterns, raising new questions about efficiency and economic implications^9,10^. These changes highlight the need for computational methods to accurately triage billable messages in accordance with policies, emphasizing clinical decision-making complexity rather than solely the time spent. This systematic review examines the computational use of SM data to enhance patient engagement, optimizing patient–provider communication, and reducing clinician burnout.

Recent advancements in computational tools and applications have shown significant potential to enhance the functionality of SM systems. NLP models have been employed to automate message classification, urgency triage, and the identification of medical decision complexity to improve clinical workflow efficiency^21,23,28–30,33,39,41^. Additionally, applications have been developed to extract social determinants of health (SDOH) from messages, revealing prevalent unmet needs of patients such as financial strain, food insecurity, and housing instability^33,42^. While BERT-based approaches have demonstrated high performance in multi-class classification, traditional ML models remain valuable in settings requiring interpretability and lower computational costs. Despite high model performance, challenges persist, particularly with class imbalances in SM datasets, which reduce algorithm sensitivity to underrepresented categories such as urgent messages and SDOH-related concerns. Moreover, the generalizability of existing tools is constrained by their development within a single healthcare institution, specialty, or patient demographic, limiting their applicability across diverse clinical settings. Since a single institution alone cannot generate sufficient evidence to fully address these limitations, future research should diversify datasets to improve model robustness and interoperability across varied patient populations^46^. Transfer learning and federated learning approaches could enhance model adaptability across institutions while preserving data privacy^47^. Furthermore, the integration of explainable AI methodologies has potential to enhance transparency and interpretability in clinical decision support, particularly for tasks with direct implications for patient care. Additionally, optimizing algorithmic trade-offs—such as maximizing recall for extracting social determinants and mental health indicators while maintaining high precision in triage systems—will be crucial for aligning computational advancements with practical clinical applications.

Beyond computational advancements, our literature screening revealed that some studies have developed theoretical frameworks based on SM corpus to support patient–provider communication. These frameworks complement computational methods in automated message processing, literacy profiling, and assessing patient engagement. One example is a theory-driven digital patient engagement model that established two robust sets of engagement measures, prioritizing factors that influence online patient interactions and using expert-reviewed data to validate engagement metrics^48^. This framework provides a structured approach for evaluating how patients interact with SM platforms, and may enhance prediction performance of computational models to support automated triage and response prioritization. Another study addressed EHR message design by integrating cognitive and behavioral science theories to enhance patient portal SM comprehension^49^. By evaluating different message formats, the study identified strategies to improve patient understanding, particularly among older adults, providing insights to inform computational developments on message simplification, readability prediction, and personalized messaging models. By integrating these frameworks with computational applications, future research could enhance clinical AI applications, ensuring that EHR systems design and implementation align with established theories of patient communication and engagement. As AI-based communication tools become increasingly embedded in clinical workflows, aligning application design with social, psychological, and behavioral theories is critical^50,51^. These frameworks provide structured guidance to bridge communication gaps, particularly in telehealth settings where engagement dynamics have evolved significantly over the past decades. Moreover, a major challenge in AI-driven messaging tools is their lack of interpretability, as many function as ‘black boxes’ with unclear decision-making processes. Theoretical frameworks contribute to greater transparency and ethical grounding by offering structured frameworks that make communication patterns more understandable and explainable. Therefore, integrating ethical guidelines, social-behavioral theories, and human-centered design principles is crucial to improving the explainability and trustworthiness of computational applications in patient–provider communication^52^.

Generative AI (GenAI) and Large Language Model (LLM) are gaining more attention in healthcare, particularly in the management of in-basket messages^53,53–55^. Recent studies have evaluated the effectiveness of GenAI-generated responses compared to those written by physicians. Research indicates that while healthcare providers often prefer human-generated messages due to their higher readability and empathy levels^56–58^, patients are generally satisfied with GenAI responses^59^. This highlights a potential acceptance of GenAI in patient interactions, depending on the context and the expectations set for communication quality. Further investigation into GenAI has focused on its ability to handle inquiries of varying complexity^60^. Advanced prompt engineering techniques and survey studies have been employed to gauge physicians’ perceptions regarding the applicability and feasibility of using GenAI for message drafting^61,62^. Additionally, pilot studies and quality improvement projects are actively exploring the practical implementation of GenAI in clinical settings^55,63^. These preliminary findings highlight the need for ongoing advancements in GenAI capabilities, particularly in handling complex messages and enhancing readability, while prioritizing patient privacy to align with the principles of meaningful use^1,2^. Our review of existing computational applications can provide valuable guidelines to inform the development of GenAI, ensuring alignment with clinical needs and patient-centered care principles. Addressing ethical considerations—including transparency in AI decision-making and safeguarding patient trust—will be critical for the responsible adoption of AI in patient–provider communication.

## Conclusion

Advancements in the computational use of secure messaging (SM) data have contributed to the evolution of SM systems, enhancing message classification, urgency triage, and patient–provider communication. However, challenges remain in ensuring model adaptability, fairness, and seamless clinical integration. To refine AI-driven solutions, a deeper synergy between computational applications and communication theories is needed. In particular, as GenAI becomes more prevalent, it is essential to leverage its capabilities to handle complex messages and enhance readability while ensuring patient privacy, ethical integrity, and alignment with e-visit billing policies. Furthermore, rigorous assessment is required to evaluate how these technologies reshape clinical workflows, ensuring they support, rather than disrupt, patient–provider interactions. Moving forward, interdisciplinary collaboration will be key to bridging technical and theoretical gaps to develop policies that balance automation with clinical oversight. By aligning computational innovation with the complex nature of patient–provider communication, SM systems can better support efficient, equitable, and high-quality patient care.

## Data Availability

The data analyzed in this study were derived from previously published literature. Synthesized data supporting the findings of this review are available from the corresponding author upon request.

## Notes

### Competing Interest Statement

The authors have declared no competing interest.

### Funding Statement

This study did not receive any funding.

